# Efficient control of IL-6, CRP and Ferritin in Covid-19 patients with two variants of Beta-1,3-1,6 glucans in combination, within 15 days in an open-label prospective randomized clinical trial

**DOI:** 10.1101/2021.12.14.21267778

**Authors:** Subramanian Pushkala, Sudha Seshayyan, Ethirajan Theranirajan, Doraisamy Sudhakar, Kadalraja Raghavan, Vidyasagar Devaprasad Dedeepiya, Nobunao Ikewaki, Masaru Iwasaki, Senthilkumar Preethy, Samuel JK Abraham

## Abstract

**Background:** Several biomarkers, including C-reactive protein (CRP), ferritin, fibrinogen, D-dimer, and Interleukin-6 (IL-6), are established predictors of disease severity and respiratory failure in COVID-19 patients.

**Objective:** In this randomized clinical study, we evaluated the efficiency of the combination of two variants’ AFO-202 and N-163 strains of Aureobasidium Pullulans produced 1,3-1,6 beta glucans in comparison with the control arm on these biomarkers in COVID-19 patients.

**Methods:** A total of 40 RT-PCR positive Covid-19 patients divided into two groups: i. control (n=22) – Standard treatment; ii. (n =18) – Standard treatment + combination of AFO-202 and N-163 beta glucans for 15 days.

**Results:** The treatment group showed decrease in CRP level compared to control. At day 7, CRP reduced to 5.53 ± 8.21 mg/L in the treatment group vs. 4.91 ± 12.54 mg/L in control (p = 0.98) (95% CI: -34.40 to 35.14). By day 15, CRP continued to be decreased at 5.42 ± 10.41 mg/L in the treatment group but it increased to 14.0 ± 37.16 mg/L in control (p = 0.52) (95% CI: -37.65 to 19.40). IL-6 levels significantly decreased in the treatment group on day 7 (p = 0.03) but the difference was not significant by day 15 (p = 0.30). Ferritin levels in the treatment group decreased from 560.58 ± 537.30 ng/mL to 127.51 ± 215.91 ng/mL by day 15, while in control increased (p = 0.98). D-Dimer level decreased in the treatment group by day 15 but was not significantly different from control (p = 0.56).

**Conclusion:** These results suggest that 15-day co-supplementation with AFO-202 and N-163 beta-glucans effectively controlled CRP, ferritin, and IL-6 in COVID-19 patients. Further research is warranted to investigate the potential of this supplement as a treatment adjunct, especially in vulnerable populations facing emerging SARS-CoV-2 variants.

## Introduction

COVID-19, caused by the SARS-CoV-2 virus, has led to a global health crisis with severe respiratory implications. Various biomarkers, such as C-reactive protein (CRP), ferritin, fibrinogen, D-dimer, and interleukin-6 (IL-6), are well-recognized as indicators of disease severity and predictors of respiratory failure in affected patients. [1]. These markers are routinely assessed upon initial diagnosis to guide treatment decisions. Various therapeutic strategies were explored for COVID-19, including immunomodulators, corticosteroids, statins, angiotensin pathway modulators, macrolides, and hydroxychloroquine. However, many of these approaches are often accompanied by undesirable side effects. With the availability of COVID-19 vaccines, adjuvants can be considered to address potential limitations such as low immunogenicity and suboptimal protective immunity [2]. Beta-glucans, particularly β-1,3-glucans, are natural immunomodulators with over 20,000 studies supporting their biological efficacy. Structurally classified as pathogen-associated molecular patterns (PAMPs), they activate immune cells through pattern Recognition Receptors (PRRs) like Dectin-1, CR3, and Toll-2 [3]. Beta-glucans have been reported to exert beneficial effects on metabolic and gastrointestinal health, reducing blood glucose, cholesterol, and aiding in conditions like metabolic syndrome and cardiovascular diseases [4]. They also have anti-infective properties against pathogens such as *Leishmania*, *Candida*, and *Staphylococcus* [5]. Yeast-derived beta-glucans, such as those from *Aureobasidium pullulans*, differ structurally and functionally [6]. The AFO-202 strain of *A.pullulans* produced beta-glucan has been reported in pre-clinical and clinical studies to be a metabolic regulator and enhances the immune response by stimulating anti-inflammatory cytokines and reducing pro-inflammatory cytokines, while N-163 strain produced beta-glucan is an immune-modulator and has anti-inflammatory and antifibrotic effects [7–9]. Unlike traditional beta-glucans that require extraction and have solubility challenges, *A. pullulans*-derived beta-glucans are water-soluble, simplifying oral administration. These beta-glucans can be explored as potential treatments and preventative measures for COVID-19 [10, 11]. Previous research showed promise for beta-glucans produced by the AFO-202 and N-163 strains of Aureobasidium pullulans in managing COVID-19 severity markers over 30 days [12]. In that study, there was beneficial effects reported in terms of sustained of erythrocyte sedimentation rate (ESR), D-Dimer, IL-6, and ferritin expression for up to 30 days. In the control groups, there was an initial decrease followed by an increase in the expression of these molecules. Effective immune enhancement and modulation were observed as a decrease in the neutrophil to lymphocyte ratio (NLR) with an increase in the lymphocyte-to-CRP ratio (LCR) and leucocyte-to-C-reactive protein ratio (LeCR).

The present study specifically investigates the effects of the combination of these beta-glucans on COVID-19 patients in the earlier stages of the illness (7 and 15 days).

## Materials and Methods

The present study was an open label, prospective, comparative, two-arm randomized pilot clinical study conducted between October 2021 and March 2022.

This study was designed as a pilot clinical trial to evaluate the effects of a food supplement in COVID-19 patients. Based on preliminary data and feasibility, the target sample size was set at 100 participants to provide an initial understanding of the intervention’s efficacy and safety.

The study aimed to enroll 100 participants; however, only 40 subjects were ultimately enrolled (Figure 1). Participants were recruited from patients presenting to the hospital following an initial triage assessment. During this process, 40 participants were randomized into the study after meeting the eligibility criteria and agreeing to participate. Recruitment challenges were compounded by the study’s focus on treating COVID-19 with a food supplement, which, despite being non-invasive, led to hesitancy among patients unfamiliar with the novel approach. Additionally, the urgency of initiating treatment during the early stages of infection further constrained the recruitment process, resulting in a smaller sample size than initially planned.

**Figure 1:**
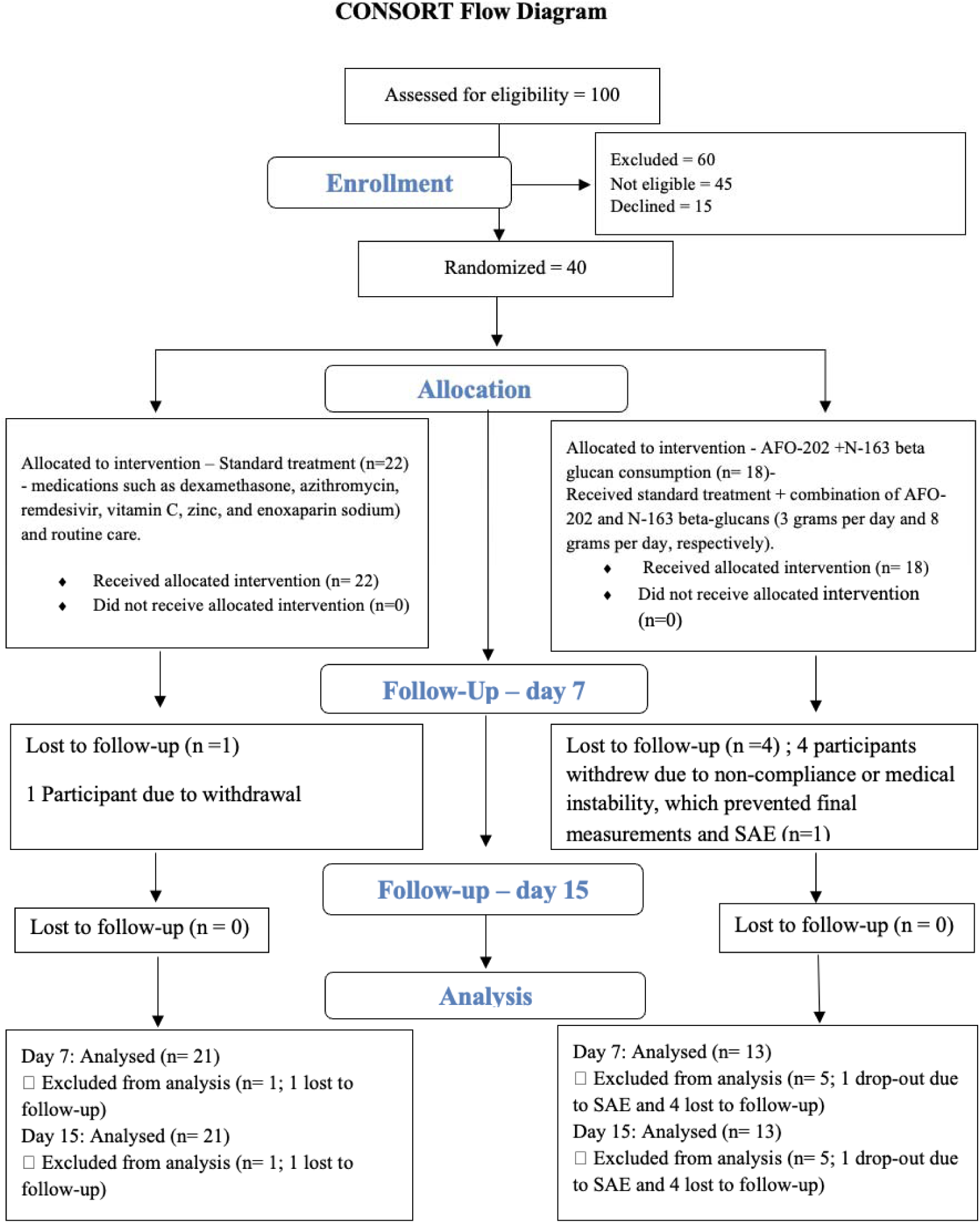
CONSORT flow diagram of the trial

The study included adult subjects aged 18-70 years (inclusive) diagnosed with SARS-CoV-2 infection via RT-PCR testing. Both genders and individuals with or without co-morbidities were eligible for enrollment, provided hospitalization was required. Patients with severe COVID-19 requiring intensive care, children, and pregnant women were excluded. Written informed consent was obtained from all participants.

Two groups (Gr.) were assigned by computer generated randomization:

- **Group 1 (Control):** Patients received standard care, comprising dexamethasone, azithromycin, remdesivir, vitamin C, zinc, and enoxaparin sodium. In addition, routine medications for pre-existing comorbidities and supportive care measures were administered as required.
- Group 2 (Treatment): Patients received standard treatment (as described in Group 1) plus a combination of AFO-202 beta-glucan at a dose of 3 grams per day (administered as 1.0 gram granules in a sachet with each meal x 3 times per day) and N-163 beta-glucan at a dose of 8 grams per day (provided in a sachet with one meal per day).

We present the CONSORT flow diagram (Figure 1) detailing the participant enrollment process. The diagram summarizes the reasons for loss to follow-up, with 5 participants lost in the intervention group and 1 in the control group. These dropout rates may have influenced the study outcomes.

The primary outcome measures of this clinical trial assessed key immunological and clinical parameters. Immunological markers, including IL-6 were monitored to evaluate the immune response. Hospitalization parameters, such as mortality, duration of hospital stay, and the need for oxygen or life-support interventions, were assessed. Comprehensive blood tests was conducted, including complete blood count (CBC), D-Dimer, C-reactive protein (CRP), erythrocyte sedimentation rate (ESR), fasting blood glucose (FBG), post-prandial blood glucose (PPG), HbA1c, lipid profile, liver function tests, and ferritin levels. The secondary outcome measures were evaluating COVID-19 clinical symptoms, recording the time taken for improvement, complete recovery, and any recurrence of symptoms such as pyrexia, respiratory distress, cephalic pain (headache), and malaise. These outcomes were measured at three key time points: Day 0 (baseline), Day 7, and Day 15.

### Statistical analysis

Statistical analysis was performed using Prism10 software. Continuous, normally distributed data are presented as mean ± standard deviation. Unpaired t-tests were employed for comparisons:. A p-value of less than 0.05 was considered statistically significant.

## Results

The study initially aimed to enroll 100 evaluable subjects; however, only 40 eligible patients were ultimately enrolled. Of these, 22 were assigned to the control group and 18 to the AFO-202+N163 treatment group (Figure 1). Notably, treatment group had a higher proportion of patients with diabetes mellitus (DM), which is known to influence the severity of SARS-CoV-2 infection (Table 1). Four patients in treatment group and one in control group withdrew from the study. Two patients in treatment group required mechanical ventilation and intensive care unit (ICU) admission, with one succumbing to the illness seven days later.

**Table 1:** Baseline characteristics of study subjects.

| **Characteristic** | **Control Group**<br>**(n=22)** | **Treatment Group**<br>**(n=18)** |
| --- | --- | --- |
| **Age Range (years)** |  |  |
| 18-25 | 0 (0%) | 1 (5.6%) |
| 26-30 | 5 (22.7%) | 3 (16.7%) |
| 31-35 | 2 (9.1%) | 0 (0%) |
| 36-40 | 4 (18.2%) | 4 (22.2%) |
| 41-45 | 3 (13.6%) | 2 (11.1%) |
| 46-50 | 4 (18.2%) | 6 (33.3%) |
| 51-55 | 2 (9.1%) | 1 (5.6%) |
| 56-60 | 1 (4.5%) | 1 (5.6%) |
| 61-65 | 1 (4.5%) | 0 (0%) |
| 66-70 | 0 (0%) | 0 (0%) |
| **Gender** |  |  |
| Male | 13 (59.1%) | 9 (50%) |
| Female | 9 (40.9%) | 9 (50%) |
| **Co-morbidities** |  |  |
| Diabetes Mellitus | 8 (36.4%) | 8 (44.4%) |
| Hypertension | 5 (22.7%) | 3 (16.7%) |
| Other Co-morbidities | 1 (4.5%) | 1 (5.6%) |
| None | 8 (36.4%) | 6 (33.3%) |
| **BMI (Mean ± SD)** | 28.0 ± 5.4 | 27.6 ± 4.6 |
| **Blood Pressure (mm/Hg) (Mean ± SD)** | 124/80 ± 15/10 | 127/82 ± 12/8 |

Randomization achieved similar age distributions in each group, with the control group ranging from 29 to 70 years (mean ± SD = 45.04 ± 11.5 years) and the treatment group ranging from 23 to 65 years (mean ± SD = 43.0 ± 9.26 years). Baseline characteristics, including body weight, body mass index (BMI), and blood pressure, were comparable across groups. All participants presented with mild to moderate COVID-19 upon admission. Oxygen supplementation (2–8 L/min) was administered to four patients in the control group and three patients in the treatment group.

While no significant differences were observed in hospitalization duration, symptom resolution, oxygen saturation on days 7 and 15, temperature, or heart rate between the two groups, it is important to note that the four patients lost to follow-up in treatment group remained hospitalized on day 15 but did not participate in the final measurements due to reasons related to their medical condition and care. Specific reasons for their failure to follow up include non-compliance and medical instability.

We evaluated the effects of treatment on key inflammatory markers, including C-reactive protein (CRP), interleukin-6 (IL-6), ferritin, and D-Dimer (Table 2).

**Table 2:** Key findings of effects on inflammatory markers: C-reactive protein (CRP), interleukin-6 (IL-6), ferritin, and D-Dimer.

| Marker | Groups | Baseline<br>(mean ± SD) | Day 7<br>(mean ± SD) | p-value<br>(Day<br>7) | 95% CI<br>(Day 15) | Day 15<br>(mean ± SD) | p-value<br>(Day<br>15) | 95% CI<br>(Day 15) |
| --- | --- | --- | --- | --- | --- | --- | --- | --- |
| CRP<br>mg/L | Control | 33.95 ± 52.89 | 4.91 ± 12.54 | 0.36 | -34.40 to 35.14 | 14.0 ± 37.16 | 0.52 | -37.65 to 19.40 |
|  | **Treatment** | **33.95 ± 61.44** | **5.53 ± 8.21** |  |  | **5.42 ± 10.41** |  |  |
| IL-6<br>pg/mL | Control | 19.48 ± 33.62 | 10.32 ± 22.493 | 0.03 | -67.40 to -<br>2.930 | 14.23 ± 29.49 | 0.3 | -45.88 to 16.83 |
|  | **Treatment** | **25.67 ± 24.9** | **8.62 ± 13.95** |  |  | **5.50 ± 7.28** |  |  |
| Ferritin<br>ng/mL | Control | 535.24 ± 605.21 | 108.18 ± 265.72 | 0.64 | -338.2 to 531.2 | 250.40 ± 356.47 | 0.98 | -371.1 to 376.6 |
|  | **Treatment** | **560.58 ± 537.30** | **278.94 ± 326.35** |  |  | **127.51 ± 215.91** |  |  |
| D-Dimer<br>FEU/L | Control | 0.91 ± 3.10 | 0.52 ± 0.39 | 0.73 | -<br>0.8059 to 1.108 | 0.38 ± 0.45 | 0.56 | -1.592 to 2.856 |
|  | **Treatment** | **0.37 ± 0.52** | **0.65 ± 1.05** |  |  | **0.29 ± 0.25** |  |  |

Baseline values of CRP were identical for the treatment and control groups (33.95 mg/L). At day 7, CRP decreased to 5.53 ± 8.21 mg/L in the treatment group and to 4.91 ± 12.54 mg/L in the control group (p = 0.98; 95% CI: -34.40 to 35.14). By day 15, CRP levels decreased to 5.42 ± 10.41 mg/L in the treatment group and 14.0 ± 37.16 mg/L in the control group (p = 0.52; 95% CI: -37.65 to 19.40) (Figure 2).

**Figure 2:**
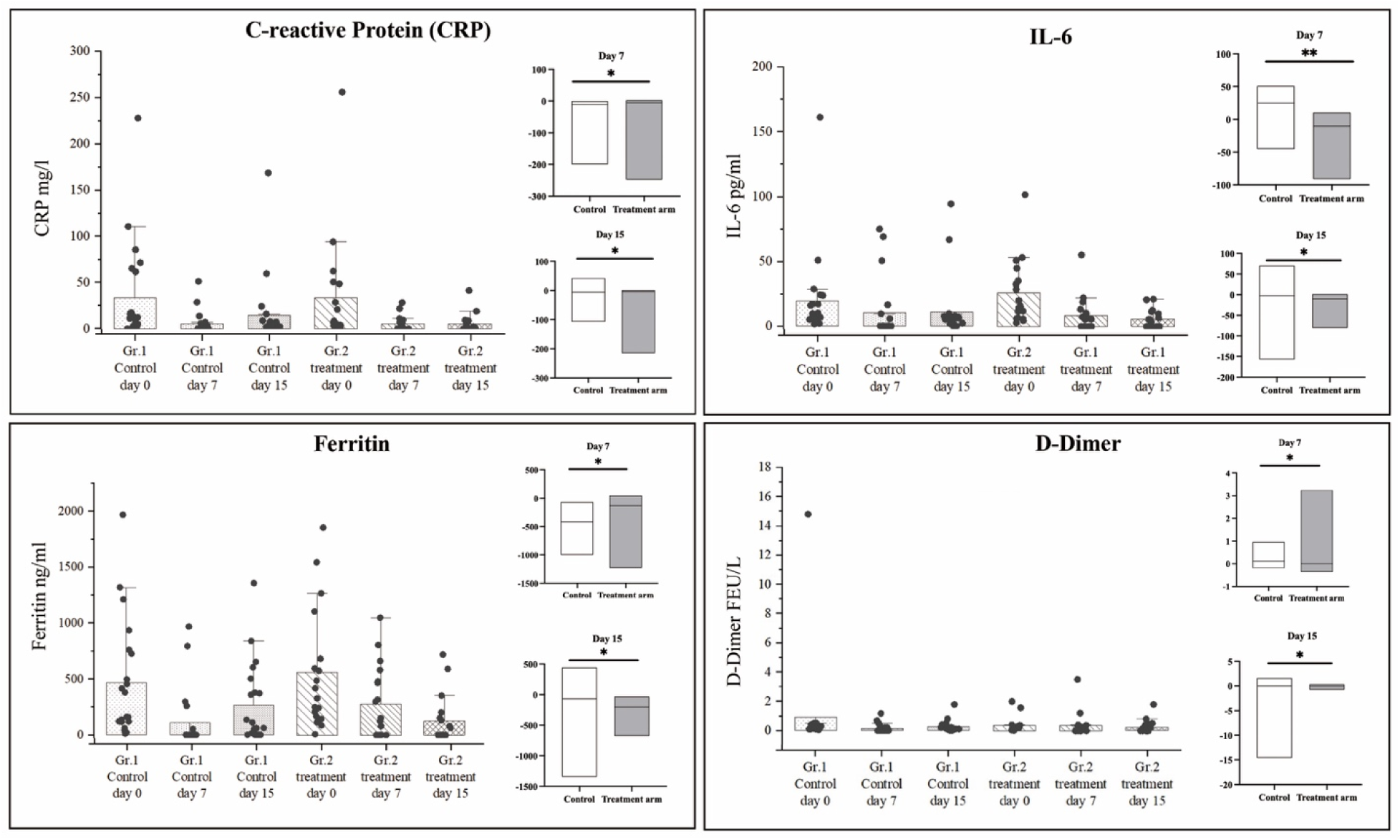
A. Decrease in CRP levels in the treatment groups compared to the control group, where levels decreased until day 7 but increased by day 15; B. Continuous decrease in IL-6 levels in the treatment groups compared to the control group; C. Decrease in Ferritin levels in the treatment groups compared to the control group, where levels decreased until day 7 but increased by day 15; D. Decrease in D-Dimer levels in the treatment groups compared to the control group, where levels decreased until day 7 but increased by day 15. The difference in values between day 7 and day 15 from baseline is depicted in separate graphs for each parameter, with median values represented by the plot lines in the floating bars. (*not significant; **p-value < 0.05 indicates statistical significance)

IL-6 levels showed a significant decrease in the treatment group at day 7 (baseline: 25.67 ± 24.9 pg/mL to 8.62 ± 13.95 pg/mL; p = 0.03; 95% CI: -67.40 to -2.930). However, by day 15, the differences between groups were no longer statistically significant (p = 0.30; 95% CI: -45.88 to 16.83) (Figure 2).

Ferritin levels decreased more steadily in the treatment group (from 560.58 ± 537.30 ng/mL at baseline to 127.51 ± 215.91 ng/mL on day 15) compared to the control group, which showed an initial decrease (535.24 ± 605.21 ng/mL to 108.18 ± 265.72 ng/mL on day 7) followed by an increase on day 15 (250.40 ± 356.47 ng/mL). Despite these trends, the differences in ferritin reduction between groups were not statistically significant at either time point (day 7: p = 0.64; 95% CI: -338.2 to 531.2, and day 15: p = 0.98; 95% CI: -371.1 to 376.6) (Figure 2).

D-Dimer levels in the treatment group declined from 0.37 ± 0.51 FEU/L at baseline to 0.29 ± 0.25 mg FEU/L on day 15, while in the control group it decreased from 0.91 ± 3.10 FEU/L to 0.38 ± 0.45 FEU/L. These changes were also not statistically significant (day 7: p = 0.73; 95% CI: -0.8059 to 1.108; day 15: p = 0.56; 95% CI: -1.592 to 2.856) (Figure 2).

HbA1c and IgA levels remained relatively stable across both groups, with no significant changes observed from baseline to day 15.

### Serious Adverse Events

Two patients in the treatment group required mechanical ventilation and admission to the intensive care unit (ICU), with one patient unfortunately succumbing to the illness seven days later. It is important to note that the adverse event was attributed to COVID-19-related complications and was determined to be unrelated to the food supplement administered as part of the intervention.

### Dropout Rates

The dropout rate was notably higher in the intervention group (n=5) compared to the control group (n=1). This significant difference in retention may have affected the overall study results and warrants further consideration in the interpretation of the findings.

## Discussion

The current study investigated the effects of the AFO-202 and N-163 beta-glucan combination in the acute phase of COVID-19. Considering the need for rapid intervention against the rapidly progressing SARS-CoV-2 pathogenesis, the combination of AFO-202 and N-163 beta-glucan from the previous study [12] was tested for a shorter duration of 7 and 15 days.

The treatment group showed evidence of a decrease of biomarkers IL-6, CRP, Ferritin and D-Dimer levels at day 7 which sustained till day 15 in the treatment group while some of the markers such as Ferritin decreased on day 7 but then increased on day 15 in the control group which suggests the more potent anti-inflammatory effects of the beta-glucans compared to the control. However, the changes were not statistically significant, making the findings warrant cautious interpretation but nevertheless these results confirm the safety of these beta-glucans as an adjuvant in COVID-19. The decrease observed in the control group may also be likely part of the natural course of COVID-19 and associated with immunologic responses or medication-related reductions in viral load, rather than solely due to supportive care received during hospitalization. In contrast, the treatment group maintained beneficial effects even after discharge.

SARS-CoV-2 infection triggers a cytokine storm, an uncontrolled release of inflammatory mediators leading to tissue damage [1]. Elevated levels of interleukin-6 (IL-6) and ferritin are associated with severe COVID-19 and increased mortality [13]. Therefore, controlling inflammation and modulating the immune response are critical therapeutic strategies. In earlier studies in healthy volunteers, a combination of AFO-202 and N-163 beta-glucans demonstrated beneficial effects, including reduced D-dimer levels, improved NLR, and increased lymphocyte-to-CRP ratio (LCR) and leukocyte-to-CRP ratio (LeCR) [14]. This combination also exhibited anti-fibrotic and anti-inflammatory properties in non-alcoholic steatohepatitis (NASH) models [8]. Another earlier study showed that this beta-glucan combination significantly decreased inflammatory and hyper-immune response markers (IL-6, NLR, ESR, D-Dimer) for 30 days, while the control group displayed increased levels of these markers on day 15 [12]. These beta-glucans, having been used as adjuvants in vaccines like influenza [6], may serve as unique nutritional supplements that complement conventional COVID-19 treatments. Having earlier demonstrated with the potential to manage the cytokine storm in COVID-19 in the 30-day study the present study highlights the potential of these AFO-202 and N-163 beta-glucan combination intervention in tackling COVID-19 in the acute phase as well. [15].

This study has several limitations. Firstly, the sample size was smaller than expected; although the study aimed to recruit 100 participants, only 40 were ultimately enrolled due to challenges in recruitment during the COVID-19 pandemic. These challenges included hesitancy toward novel treatments and logistical difficulties. Secondly, the study’s follow-up duration was shortened from the intended 60 days to 15 days, as many participants were unwilling to continue the supplement post-discharge. This limited the ability to assess long-term outcomes and the sustained effects of the treatment beyond the acute phase of infection. The next limitation is the lack of a placebo-control group. Finally, the dropout rates related to patient follow-up post-discharge represent another limitation, as it restricted further data collection in the later stages of recovery. These factors suggest that while the findings are promising, they should be interpreted with caution, and larger, longer-term studies are necessary to confirm these results.

Further, fasting blood glucose levels have been associated with the severity of SARS-CoV-2 infection [16]. While our study did not show significant differences in clinical outcomes such as hospitalization duration or inflammatory markers between the groups, the presence of a higher proportion of DM patients in treatment group (Table 1) could potentially have influenced the results. DM is known to exacerbate inflammation and other COVID-19-related complications, which may have contributed to greater variability in the response to treatment within the treatment group. Previous studies have demonstrated that AFO-202 beta-glucan exhibits metabolic regulatory effects, including a reduction in HbA1c levels and regulation of dyslipidemia in human clinical studies [17,18]. Additionally, when combined with N-163 beta-glucan in a NASH mouse model, it showed further metabolic regulation and immune modulation [5]. Therefore, it is plausible that the beta-glucan treatment, particularly its metabolic-regulating properties, may have contributed to more pronounced immune modulation in patients with diabetes mellitus (DM). However, the current study was not designed to analyze specific subgroups, and we did not perform an analysis stratified by comorbidities like DM. Future studies focusing on DM patients alone could provide valuable insights into whether beta-glucan supplementation offers additional benefits in managing COVID-19 among patients with diabetes. We recommend this as an area for further research.

## Conclusion

The present study demonstrates that co-supplementation with AFO-202 and N-163 beta-glucans from *Aureobasidium pullulans* modulated key biomarkers associated with COVID-19 disease severity and mortality, including interleukin-6 (IL-6), C-reactive protein (CRP), and ferritin. These markers remained decreased in the treatment group compared to the control group receiving standard care alone, at both the 7-day and 15-day follow-up points. These findings support the potential of these beta-glucans as orally administered adjunctive therapy for COVID-19 treatment and prevention. However, further validation through larger, multicenter clinical trials is necessary.

## Data Availability

All data produced in the present work are contained in the manuscript

## Acknowledgements

The authors thank

1. The Government of Japan and the Prefectural Government of Yamanashi for a special loan and M/s Yamanashi Chuo Bank for processing the transactions.
2. Dr. Ragaroobine, Mr. Mathaiyan Rajmohan of NCRM, Dr. Subbiah Sriraam, Ms.Aditi Ramesh and staff of Aurous HealthCare for their assistance with the regulatory procedures, documentation, logistics and data collection of the study.
3. Mr. Takashi Onaka (Late) and Mr. Yashushi Onaka (Sophy Inc, Kochi, Japan), for necessary technical clarifications.
4. Mr. Yoshio Morozumi and Ms. Yoshiko Amikura; of GN Corporation, Japan for their liaison assistance with the conduct of the study; Dr. Rajappa Senthilkumar and Dr. Koji Ichiyama of GN Corporation, Japan for technical assistance.

## Declarations

### Ethics Approval

The study was registered in Clinical trials registry of India, CTRI/2021/10/037380. The study was approved by the Institutional Ethics Committee (IEC) of Madras Medical College, India on 15^th^ September, 2021.

### Availability of data and material

All data generated or analysed during this study are included in this manuscript

### Author Contribution Statement

S.U.P and S.A. contributed to conception and design of the study. E.T and D.S helped with data collection. S.A and S.P. drafted the manuscript. K.R, S.S, V.D, N.I and M.I performed critical revision of the manuscript. All the authors read, and approved the submitted version. All the authors meet the criteria for authorship as per the ICMJE criteria

